# A Survey of Electronic Health Record (EHR) Variables Associated with Postoperative Delirium: A Systematic Review

**DOI:** 10.1101/2020.05.28.20116251

**Authors:** Matthew M Ruppert, Haleh Hashemighouchani, Emel Bihorac, Seth Williams, Laura Velez, Julie S Cupka, Tezcan Ozrazgat-Baslanti, Parisa Rashidi, Azra Bihorac

**Affiliations:** Department of Medicine, University of Florida, Gainesville, FL, USA; Precision and Intelligent Systems in Medicine (Prisma^P^), University of Florida, Gainesville, FL, USA; Department of Biomedical Engineering, University of Florida, Gainesville, FL, USA

## Abstract

**Introduction:** Delirium is a common post-operative complication in critically ill patients, displaying transient changes in consciousness, inattention, awareness, and organized thought. Not a lot is known about the specific causes of this condition, as it is a complex physiologic state that is currently being unraveled to determine any correlation with imbalances in homeostasis.

**Objective:** The aim of this systematic review is to report on and summarize risk factors associated with the development of postoperative delirium in critically ill adult patients.

**Methods:** This systematic review assessed studies reporting on risk factors for postoperative delirium in critically ill patients. PubMed, PsycINFO, and CINAHL databases were searched for studies. Observational or interventional studies reviewing predictors for postoperative delirium in delirious versus non-delirious patients were included.

**Results:** Fifty potential risk factors were identified and divided into eight subgroups. The significance of a specific risk factor for postoperative delirium was found to depend on the patient population in question, but consistently cited significant risk factors across cohorts included high mortality risk, abnormal laboratory values, and use of vasopressors, analgesics, thiopentones, propofol, and benzodiazepines. There was not enough evidence found to definitively state the significance of type of surgery being performed or cognitive impairment on development of this condition.

**Conclusion:** Several risk factors were found to be significantly associated with the development of postoperative delirium, including high risk of mortality, specific medication use, and abnormal laboratory values. Further research needs to be performed to fully define the importance of these individual risk factors across broad critical care population.

## INTRODUCTION

Postoperative delirium (POD) is a dangerous complication that is defined as a neuropsychiatric disorder associated with unstable levels of consciousness, disorganized thought, inattention, and loss of awareness [1]. This common illness results in increased rates of morbidity and mortality, extended hospital stays, and reduced quality of life [2]. In the United States, the healthcare costs of delirium are comparable to that of diabetes and mainly lies among older adults [3].

Given the prevalence and epidemiological complexity of POD, it is imperative to engage in an in-depth investigation of the factors that influence whether a patient will develop delirium after surgery [4–6]. While 30–40% of POD cases are preventable, delirium is vastly underdiagnosed within the intensive care unit (ICU) [7]. The diagnostic tools most commonly used to assess for delirium in the ICU include the Delirium Symptom Interview, Saskatoon Delirium Checklist, Delirium Rating Scale-Revised Version (DRS-R-98), Memorial Delirium Assessment Scale (MDAS), Confusion Assessment Method (CAM), CAM-ICU, and Clinical Assessment of Confusion-A and –B (CAC-A/B) [8]. These assessment methods are useful when performed correctly but unfortunately that is not always the case. Several nurses may chart CAM results based on unit protocols to deliver optimal patient care, which may or may not reflect a true CAM status for the patient [9]. Detection of delirium is not the only problem at hand, as there is currently no proven treatment for delirium [10–12]. As a result, prevention of delirium is the most effective mode of treatment [7]. It may be pragmatic to focus biomedical delirium research on a standardized, automated detection system that can assess patients for all potential risk factors. The purpose of this review is to examine currently available literature and summarize delirium-associated risk factors for post-operative adult ICU patients.

## METHODS

PubMed, PsycINFO, and CINAHL databases were used to search four different sets of key words. Each database was initially searched with the following key words: 1) “Delirium AND (ICU OR intensive care unit OR critical care) AND (risk stratification OR risk assessment OR risk model)”; 2) “Delirium AND (ICU OR intensive care unit OR critical care) AND (predict*)”; 3) “Delirium AND (ICU OR intensive care unit OR critical care) AND (predict OR prediction OR predictive)”; 4) “Delirium AND (ICU OR intensive care unit OR critical care) AND (detect OR detection OR detection system)”. After obtaining a raw collection of articles, a systematic approach was used to exclude articles. In the title and abstract screening phase, articles published prior to 2012 or after 2017, articles not written in English, articles without a post-operative patient population, and duplicate articles were excluded. Then, the full text of the remaining articles were reviewed. Articles were further excluded that were not based on an observational study or clinical trial, were not reporting delirious and non-delirious subject data, did not have applicable data, analysis was based on secondary source data, did not have a control group, contained undesired population characteristics, did not contain original data, or the full text was unavailable. The full text review was performed with guidance from the Mixed Methods Appraisal Tool (MMAT) to identify the articles meeting all of our inclusion criteria [13]. The Preferred Reporting Items for Systematic Reviews and Meta-Analyses (PRISMA) checklist guidelines were followed when reporting our systematic review [14] (Supplementary Table 1).

Data from the final 27 articles was then extracted into a spreadsheet containing the sample size, number of control patients, number of delirium patients, delirium diagnostic method, examined data elements, statistical tests, and probabilities of each element associated with delirium. An extraction sheet provided a method to perform a descriptive analysis for the compiled studies so that reliability and validity could be measured. The data was then used to categorize all identified delirium risk factors into generalized subgroups.

## RESULTS

### Literature search results & outcome

The four search strings used in PubMed, PsycINFO, and CINAHL returned a total of 1,678 articles. After the removal of 772 duplicates, 956 items remained. The first phase of elimination removed a total of 864 articles reducing the number of articles to 92 to be fully reviewed. The full-text review of these articles revealed a total of 27 publications to be used in determining predisposing elements associated with postoperative delirium in critically ill adults (Figure 1).

**Figure 1.**
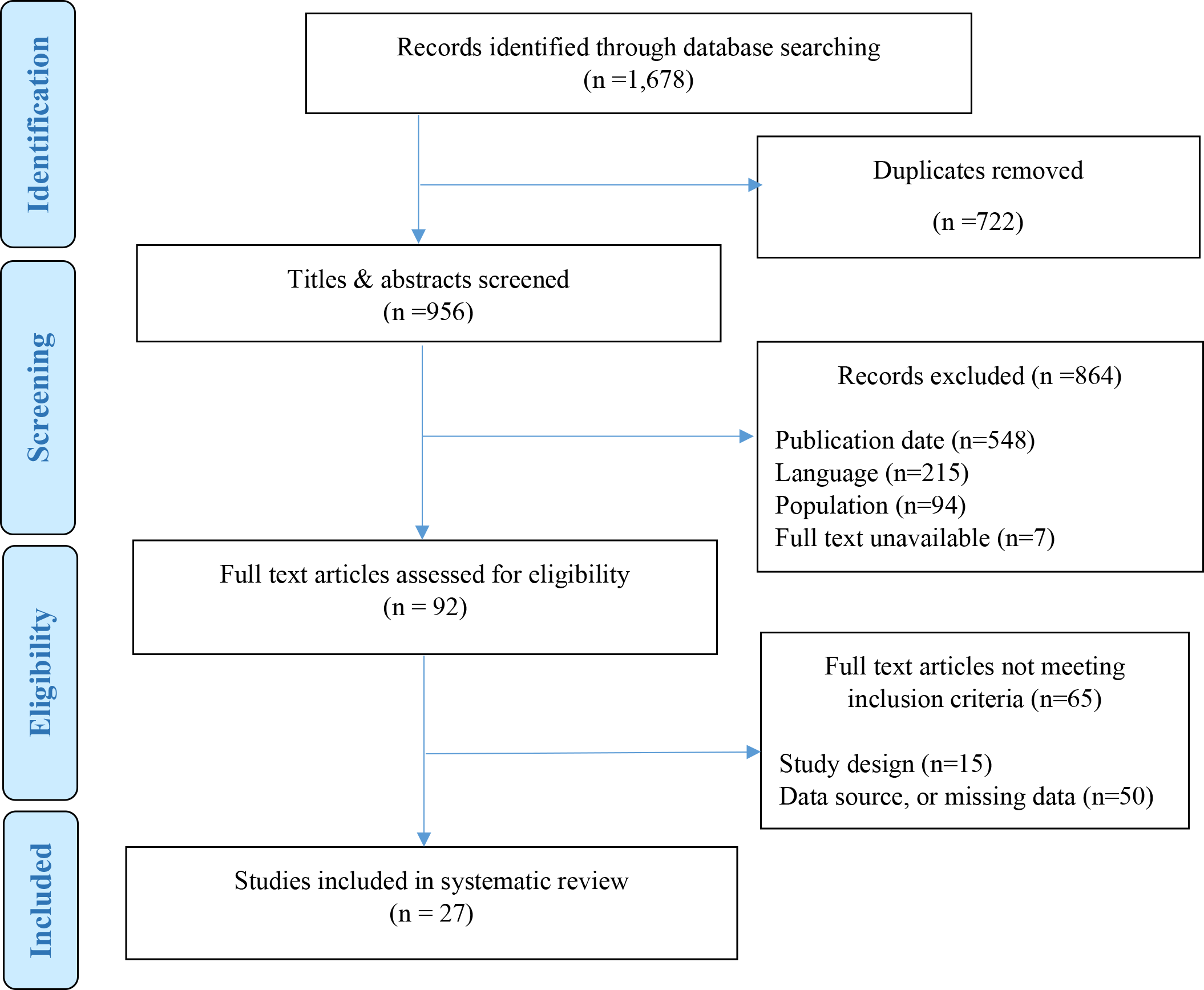
PRISMA Record Screening Flow chart.

### Study characteristics

Both prospective and retrospective cohorts were used for data collection. The sample sizes ranged from 23 to 768 subjects. Fifty potential risk factors for postoperative delirium were identified during data extraction, which were further divided into eight subgroups (Table 1).

**Table 1.** Potential Risk Factors for Postoperative Delirium.

|  |  |
| --- | --- |
| Demographic Factors | Age<br>Sex<br>Smoking/nicotine abuse<br>History of alcohol consumption<br>Sleep |
| Medications | Preoperative anti-depressants<br>Anesthesia type<br>Inotropic agents<br>Vasopressors<br>Analgesics |
| Physiologic Variables | Low serum albumin<br>High bilirubin<br>Bispectral index (BIS)<br>Cardiac function<br>High C-reactive protein (CRP)<br>Low hematocrit<br>Low hemoglobin<br>Hypertension<br>Hypoxia<br>Decreased blood proteins<br>Decreased red blood cell count (RBC)<br>Increased white blood cell count (WBC)<br>Sodium<br>High creatinine |
| Comorbidities and Complications | Atrial fibrillation<br>History of stroke<br>History of cerebrovascular disease<br>Cognitive impairment<br>Impaired executive functions<br>Congestive heart failure (CHF)<br>Chronic obstructive pulmonary disease (COPD)<br>Diabetes mellitus (DM)<br>Intraoperative blood loss |
| Medical Interventions | Type of surgery<br>Type of anesthesia<br>Blood transfusion<br>Cardiopulmonary bypass (CPB)<br>Ventilator use<br>Non-invasive ventilator use |
|  | Renal replacement therapy (RRT)<br>Use of intra-aortic balloon pump (IABP) |
| Scoring Systems | High Acute Physiologic Assessment and Chronic Health Evaluation II score (APACHE II)<br>Low Mini-Mental State Exam Score (MMSE)<br>High Charlson comorbidity index<br>High EuroSCORE<br>High American Society of Anesthesiologists Physical Status Classification System score (ASA PS) |
| Length of Treatments | Duration of surgery<br>Duration of mechanical ventilation<br>Duration of anesthesia<br>Duration of cardiopulmonary bypass time |

### Demographic factors

Age was examined as a predictor for delirium in 26 of the 27 papers reviewed. Six of the studies investigated age by using cutoffs as a risk factor. Four of these found significant cutoffs: one at 50 years [15], one at 60 years [16], one at 65 years [17], and one at 70 years-old [18]. The two studies that did not find significance used 65 years [19] and 80 years-old [20] as cutoffs. Of the 21 studies that examined sex as a predictor for delirium, only one study found significance reporting that females are more likely to develop the condition [21]. Seven studies investigated smoking status as a predictor for delirium, three of which found smoking to be a significant risk factor. Of those finding significance, two had a population of post-operative cardiac patients[20, 22], and one examined general surgery patients[23].

Seven studies assessed a history of alcohol consumption as a predictor for delirium, three of which found a significant association between the two variables. The populations in the studies where alcohol is a significant predictor were comprised of liver transplant [24], severe burn [15], and major general surgery patients [23] (Table 2).

**Table 2.** Correlation Between Demographic Predictors and Postoperative Delirium.

| Variable | Total Studies | $p \leq 0.005$ | $p < 0.05$ | $p \geq 0.05$ |
| --- | --- | --- | --- | --- |
| Age | 26 | 6 | 8 | 12 |
| Sex | 21 | 0 | 1 | 20 |
| Tobacco Use | 7 | 1 | 2 | 4 |
| Alcohol Use | 7 | 2 | 1 | 4 |
| Sleep | 5 | 1 | 2 | 2 |

### Medications

Of the 27 studies analyzed, 13 examined pharmacological risk factors in relation to postoperative delirium. It was shown in one prospective study that the number of concomitant medications, regardless of the combination, is a significant risk factor for developing the condition [23]. Forty-three percent of delirious patients vs twenty-three percent of non-delirious patients had six or more concomitant medications [23]. Another study found that patients using a calcium channel blocker were more likely to develop delirium [22]. Among studies on analgesics and vasopressors, there was a consensus that both are significant risk factors for delirium [17, 19, 25–27]. The doses of midazolam [28] and propofol [16] during surgery have been reported as significant risk factors for developing delirium; however, a more recent study found midazolam use not to be a significant risk factor [16]. In a prospective observational study, the anesthetic thiopentone demonstrated an eight-fold-higher risk for patients developing delirium when compared to propofol [19]. Preoperative anti-depressant use was shown to be a risk factor for delirium in only one the three studies investigating this variable [22, 23, 29]. When anti-depressants were analyzed individually, selective serotonin reuptake inhibitors (SSRIs) were found to be significant in a cohort of postoperative cardiac patients but not amongst general surgery patients [23, 29].

Tricyclic/tetracyclic antidepressants were consistently found to be an insignificant risk factor [23, 29]. The type and duration of inotropic medications has also been reported to be associated with POD [25, 26], where dopamine infusion emerged as an independent risk factor in patients undergoing CABG [25]. Benzodiazepine use was designated as a significant risk factor; however, other hypnotics have not been found to have similar effects [23] (Table 3).

**Table 3.** Correlation Between Medication Use and Postoperative Delirium.

| Variable | Total Studies | $p \leq 0.005$ | $p < 0.05$ | $p \geq 0.05$ |
| --- | --- | --- | --- | --- |
| Anti-depressants | 3 | 1 | 0 | 2 |
| Anesthesia Type | 13 | 2 | 3 | 8 |
| Inotropic Agents | 5 | 2 | 0 | 3 |
| Vasopressors | 2 | 2 | 0 | 0 |
| Analgesics | 4 | 2 | 2 | 0 |

### Physiological Variables

Low serum albumin levels are a fairly strong delirium risk factor with three out of four studies reporting it as significant [23, 30, 31]. An elevated amount of CRP is a very strong risk factor for delirium with four of four studies finding it significant [20, 23, 31, 32]. Decreased hemoglobin appears to be a moderately strong risk factor for delirium, with six of the nine studies finding it significantly decreased amongst delirious patients [18, 23, 28, 31–33]. Another very strong risk factor is a decrease in blood proteins, with both studies investigating this factor finding a correlation coefficient below 0.005 [23, 31]. A decreased RBC count is a strong risk factor for delirium with two articles demonstrating a highly significant correlation between a decrease in RBC and delirium [31, 34]. Elevated creatinine levels are a strong risk factor for delirium with nine of twelve studies finding it significant [16, 23, 27, 28, 32, 33, 35] (Table 4).

**Table 4.** Correlation Between Physiological Variables and Postoperative Delirium.

| Variable | Total Studies | $p \leq 0.005$ | $p < 0.05$ | $p \geq 0.05$ |
| --- | --- | --- | --- | --- |
| Serum Albumin | 4 | 3 | 0 | 1 |
| Bilirubin | 3 | 1 | 0 | 2 |
| BIS | 2 | 1 | 1 | 0 |
| Cardiac Function | 5 | 1 | 1 | 3 |
| CRP | 4 | 2 | 2 | 0 |
| Low Hematocrit | 4 | 1 | 1 | 2 |
| Hemoglobin | 9 | 3 | 3 | 3 |
| Hypertension | 16 | 2 | 1 | 13 |
| Hypoxia | 5 | 3 | 0 | 2 |
| Blood Proteins | 2 | 2 | 0 | 0 |
| Decreased RBC | 2 | 2 | 0 | 0 |
| Increased WBC | 2 | 2 | 0 | 0 |
| Sodium | 4 | 1 | 0 | 3 |
| Creatinine | 12 | 7 | 2 | 3 |

### Co-morbidities and medical history

Pre-operative cognitive impairment and impaired executive functions were consistently cited as a risk factor for post-operative delirium [20, 26, 28, 36]. However, the validity of these factors as delirium predictors should be taken with a degree of skepticism because cognitive disorders such as dementia present similarly to delirium and vice versa, often leading to misdiagnosis [37]. The other historical risk factors examined in our review, such as history of stroke, did not consistently report a significant association with delirium (Table 5).

**Table 5.** Correlation Between Comorbidities, Complications, and Postoperative Delirium.

| Variable | Total Studies | $p \leq 0.005$ | $p < 0.05$ | $p \geq 0.05$ |
| --- | --- | --- | --- | --- |
| Atrial Fibrillation | 7 | 4 | 0 | 3 |
| Stroke | 6 | 1 | 2 | 3 |
| Cognitive impairment | 2 | 2 | 0 | 0 |
| Impaired executive functions | 7 | 3 | 4 | 0 |
| Congestive Heart Failure | 3 | 1 | 1 | 1 |
| COPD | 6 | 0 | 1 | 5 |
| Cerebrovascular Disease | 3 | 1 | 0 | 2 |
| Diabetes Mellitus | 15 | 2 | 3 | 10 |
| Intraoperative blood loss | 5 | 2 | 2 | 3 |

Intraoperative blood loss has also been shown as a predictor for development of POD in two population subsets: elderly patients receiving total hip arthroplasty (THA) procedures and severely burned patients undergoing early escharotomies [15, 31]. Large volume blood loss can lead to intraoperative hypotension and an increased need for blood transfusions, both of which have been shown to be predictors for POD [15, 31] (Table 5).

### Medical interventions

Five of ten studies demonstrated a significant association between greater volumes of blood transfusion and higher risk of POD [16, 18, 26, 31, 38]. Anemia and reduction of blood oxygen level could be another factor that effects neuropsychological performance by inducing transient ischemia [18]. There was no remarkable association between type of surgery and POD [36, 39] when comparing a wide range of surgery types; however, trauma patients had a higher risk of POD occurrence [36]. Two of four studies evaluating the relationship between POD and emergency surgery showed that emergent surgery has a higher risk of POD [23, 36]. The association between POD and open-heart surgery including CABG and valvular surgeries were studied by 10 investigators, four of which demonstrated significant results [20, 21, 26, 35]. Higher risk of POD was associated with more complex open-heart surgeries as a result of longer duration of surgery [35].

Exposure to CPB was found to be a risk factor for POD in two of three studies [18, 40]. CPB is known as one of the causes of intraoperative hyperoxic cerebral reperfusion which is associated with higher incidence of POD [40].

Additionally, mechanical ventilation is a significant predictor for POD [26, 29, 35]. A pilot study also demonstrated remarkable association between post-operative, non-invasive mechanical ventilation and POD, recognizing cerebral hypoxia as one of the important causes for POD [16] (Table 6).

**Table 6.** Correlation Between Medical Interventions and Postoperative Delirium.

| Variable | Total Studies | $p \leq 0.005$ | $p < 0.05$ | $p \geq 0.05$ |
| --- | --- | --- | --- | --- |
| Type of surgery |  |  |  |  |
| Open heart surgery | 10 | 1 | 3 | 7 |
| Emergent surgery | 4 | 0 | 2 | 2 |
| Amputation/Vascular | 1 | 1 | 0 | 2 |
| Trauma | 1 | 1 | 0 | 0 |
| Type of anesthesia | 13 | 2 | 3 | 8 |
| Blood transfusion | 10 | 4 | 1 | 5 |
| CPB | 3 | 0 | 2 | 1 |
| Ventilation use | 3 | 1 | 1 | 1 |
| Non-invasive ventilation use | 1 | 1 | 0 | 0 |
| RRT | 3 | 1 | 1 | 1 |
| IABP use | 2 | 0 | 0 | 2 |

### Scoring Systems

There are many scoring systems used to assess the health status of a patient. Of all the scoring systems examined to predict POD, the use of the ASA PS system was the most commonly found assessment method in the literature as a potential aid for prediction of delirium. The ASA PS is a subjective measure of preoperative comorbidities and the patient’s condition [41]. A higher ASA PS score was found to have a significant positive correlation to post-operative delirium risk in three of seven reviewed studies looking into it as a factor. The APACHE II score is another popular scoring system that is used as a postoperative delirium prediction aid. One study found that 70% of delirious patients had an APACHE II score ≥ 16

[24], and all four studies examining APACHE II as a potential risk factor found a high score to be significantly associated with postoperative delirium [24, 36, 38, 39] (Table 7).

**Table 7.** Correlation Between Scoring System and Postoperative Delirium.

| Variable | Total Studies | $p \leq 0.005$ | $p < 0.05$ | $p \geq 0.05$ |
| --- | --- | --- | --- | --- |
| APACHE II score | 4 | 4 | 0 | 0 |
| MMSE score | 6 | 3 | 1 | 2 |
| Charlson comorbidity index | 6 | 2 | 1 | 3 |
| EuroSCORE | 6 | 2 | 1 | 3 |
| ASA PS | 7 | 2 | 1 | 4 |

### Length of treatments

In the subgroup describing length of treatments as possible risk factors, long surgery duration was the most prevalent of these risk factors, with the median time for duration of surgery ranging from 72 minutes to 616 minutes. The combination of longer surgery duration entails more complex procedures, larger doses of anesthetic, and greater volumes of blood transfusion which increase exposure to hypoperfusion and systemic inflammatory response [26]. These factors can cause postoperative complications, including electrolyte disturbances, which are reported to result in POD [26].

Another potential risk factor is the time spent under mechanical ventilation. This risk factor is correlated with both duration of surgery and of anesthesia, as longer anesthetized procedures require longer mechanical ventilation [18, 34]. When patients are intubated, they are sedated by drugs which can alter brain function and cause increased risk for delirium [27]. Thusly, the longer the patient is sedated and intubated, the higher the risk of POD due to prolonged altered brain activity. One study found that patients intubated for five days or longer were 1.81 times more likely to develop POD than those intubated for four days or less, and another study estimates that extended periods of intubation may raise the prevalence of delirium in post-operative patients by 10% [24, 27].

Of note, while Liu et al. [27] reported a significant association between duration of CPB and incidence rates of POD, seven studies did not reveal CPB duration as a risk factor for POD. Only one study found that reinstituting CPB during an operation increased the risk of developing POD[18] (Table 8).

**Table 8.**
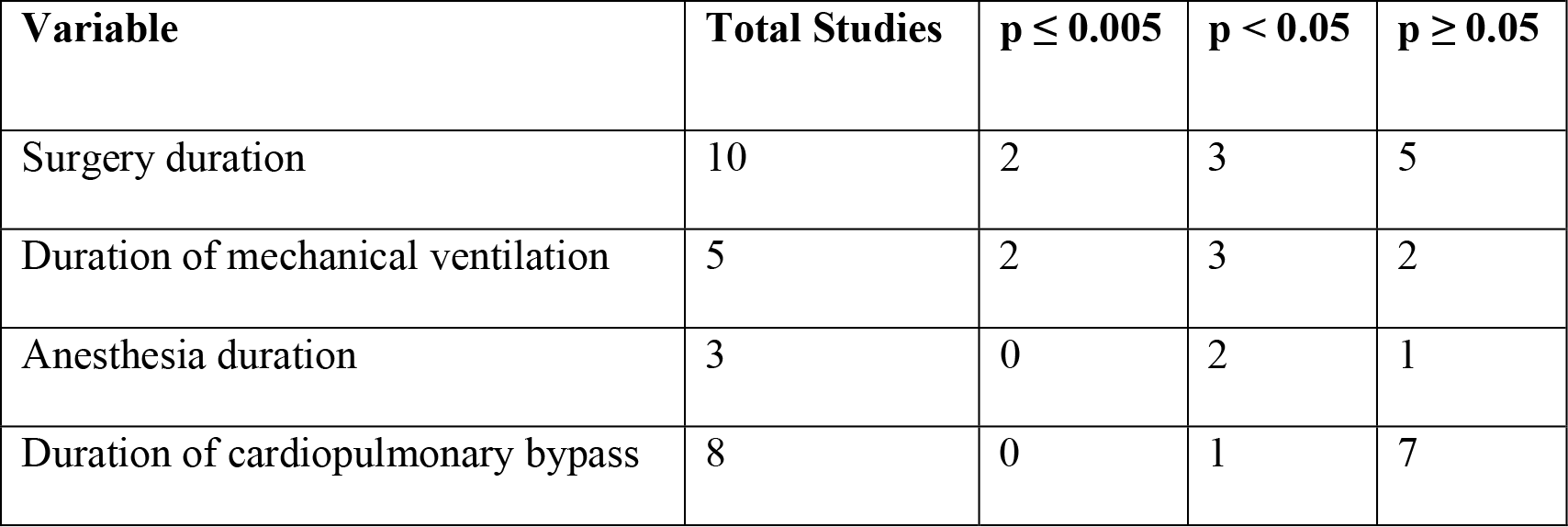
Correlation Between Duration of Interventions and Postoperative Delirium.

## DISCUSSION

Through the course of this review there were 50 potential risk factors identified, many of which were only found to be significant amongst certain populations. Medications that were consistently cited as a significant risk factor include: vasopressors, analgesics, thiopentones, propofol, and benzodiazepines. Cardiac patients were found to have the use of SSRIs as a significant risk factor for delirium. Changes in blood composition, specifically decreased RBC, increased WBC, decreased blood proteins, elevated serum creatinine, and lower hemoglobin, have been consistently correlated with increased risk of delirium. A definitive statement on the effect of surgery type on the development of delirium cannot be made, because the significantly correlated types of surgery have not been corroborated by other sources as of this publication. A high APACHE II score was overwhelming associated with a higher risk of delirium. Cognitive impairment was found to be consistently associated with increased likelihood of delirium; however, further research will need to be done to verify this association is due to delirium and not a worsening of the underlying cognitive impairment.

The studies that focused on demographical factors and their effects on delirium analyzed age, sex, tobacco use, alcohol use, and sleep. Most studies which used age as a prediction factor found significant results. However, the studies pertaining to age which showed a strong relationship between age and delirium had a larger underlying age difference between delirious and non-delirious groups and/or a small variance in age within each group compared to those that did not. The reason for the increased prevalence of delirium among older adults is believed to be the result of a combination of increased neurotransmitter disturbances [42], neuron apoptosis, and decreased cerebral blood flow[15]. The data collected suggests that the remaining prediction factors in this subgroup are not necessarily risk factors for delirium, however, these factors should provide an avenue for further research.

It is widely accepted that certain drugs are more likely to induce delirium, however it has been shown in one prospective study that that the number of concomitant medications, regardless of the combination is also a significant risk factor [23]. While among studies on analgesics and vasopressors, there was a consensus that both are significant risk factors for delirium [17, 19, 25–27]. There is a lack of information on the effects of preoperative anti-depressant use on delirium which requires more research with narrow focus. Of the three articles found on antidepressant use, only one found any significance which could potentially be explained by the way the different papers grouped anti-depressants in their analysis [22,23]. Further research should aim to discover which antidepressants influence the onset of delirium. Other pharmacological risk factors were either not reported, or were not found to be significant risk factors in most of the studies.

There was a large number of strong physiological markers that were associated with delirium. Studies investigating CRP and low serum albumin levels show that they are both fairly strong delirium risk factors with a vast majority of the studies on each reporting them as significant [12,15, 23–25]. A decrease in blood proteins is a very strong risk factor for delirium, with both studies examining it finding a correlation coefficient below 0.005 [15, 24]. Both elevated creatinine levels and decreased RBC count showed a strong correlation that they are risk factors for delirium, however, there was not a complete agreement on this ad there were studies apposing these findings.

In this subgroup, the most commonly assessed risk factor was blood transfusion. Transfusion of stored RBCs, particularly those transfused during surgery, appear to induce a peripheral inflammatory cytokine profile similar to that seen in POD [43]. Anemia and reduction of blood oxygen level could be another factor that effects neuropsychological performance by inducing transient ischemia[18]. There was no remarkable association between surgery type and POD [36, 39] when comparing a wide range of surgery types. Several studies found a significantly higher risk of POD in association with more complex open-heart surgeries as a result of longer duration of surgery[35]. Mechanical ventilation has been a significant predictor for POD[26, 29, 35], but a pilot study also demonstrated remarkable association between postoperative non-invasive mechanical ventilation and POD, recognizing cerebral hypoxia as one of the important causes for POD[29]. This study laid framework to build on and develop an understanding towards the influence of using non-invasive mechanical ventilation on the onset of delirium.

The ASA PS was the most commonly found scoring system in the literature as a potential predictor for delirium. A higher ASA PS score was found to have a significant positive correlation to post-operative delirium risk in three of seven reviewed that used it as a factor. However, the APACHE II score was also popularly noted. A patient’s APACHE II score has been found to be significantly associated with the onset of postoperative delirium [16, 29, 32, 36]. Other scoring systems were not reported to have a significant correlation with predicting delirium in postoperative patients.

Pre-operative cognitive impairment and impaired executive functions were unsurprisingly consistently reported as a significant risk factor for POD; however, the validity of these factors as a predictor for delirium should be taken the consideration that other cognitive disorders and disturbances present similarly to delirium which can provide false positive or negative delirium screenings [37]. Other risk factors examined in our evaluation did not consistently report significant association with delirium.

Intraoperative blood loss has been shown as a predictor for development of POD [15, 31]. Intraoperative hypotension and an increased need for blood transfusions can result from intraoperative blood loss; furthermore, both have been shown to be predictors for POD [15, 31]. Given this information, reducing the incidence rate and duration of intraoperative hypotension can likely improve the likelihood that a patient will not develop POD, and suggests additional risk assessment when considering administration of blood transfusions.

It has been posed that a longer surgery duration entails more complex procedures, larger doses of anesthetic, and greater volumes of blood transfusion. Which can lead to complications such as electrolyte disturbance which has been found to result in POD [18]. Systemic inflammatory response and increased need for blood transfusions have been determined to be independent risk factors and could be strongly tied to length of surgery [16, 18, 33]. Furthermore, the length of time spent under mechanical ventilation is correlated with both duration of surgery and of anesthesia, due to longer procedures requiring prolonged mechanical ventilation [18, 34]. As previously discussed, complex surgeries have many risk factors for POD, and prolonged intubation is another component to consider.

There is still much to learn about the underlying pathophysiology of delirium, making the identification of specific risk factors very difficult. Many of the factors identified in this review have significant overlap and are a result of a combination of medical procedures imposed on a variety of underlying diseases. We propose that further research into delirium associated risk factors be carried out at both a grand scale and biomolecular scale. The grand scale will allow the examination of thousands of cases of delirium and non-delirium through the analysis of electronic health record data to collect a list of all potential factors.

The biomolecular approach could shed light on the fundamental biochemical processes that underlie delirium and would be used to narrow the field of potential risk factors. The outcomes of these investigations can be used to not only identify preventative measures for delirium prevention, but potentially treatments as well.

## CONCLUSION

Several risk factors were found to be significantly associated with the development of postoperative delirium, including high risk of mortality, specific medication use, and abnormal laboratory values. Further research needs to be performed to fully define the importance of these individual risk factors across broad critical care population.

## Data Availability

NA. Systematic review paper

## References

1. van Meenen, L.C., et al., Risk prediction models for postoperative delirium: a systematic review and meta-analysis. J Am Geriatr Soc, 2014. 62(12):p.2383–90.

2. Francis, J. andW.N. Kapoor, Prognosis after hospital discharge of older medical patients with delirium. J Am Geriatr Soc, 1992. 40(6): p. 601–6.

3. Leslie, D.L., et al., One-year health care costs associated with delirium in the elderly population. Arch Intern Med, 2008. 168(1): p. 27–32.

4. Lin, W.L., Y.F. Chen, and J. Wang, Factors Associated With the Development of Delirium in Elderly Patients in Intensive Care Units. J Nurs Res, 2015. 23(4): p. 322–9.

5. Kalabalik, J., L. Brunetti, and R. El-Srougy, Intensive care unit delirium: a review of the literature. J Pharm Pract, 2014. 27(2): p. 195–207.

6. Page, V.J., et al., Effect of intravenous haloperidol on the duration of delirium and coma in critically ill patients (Hope-ICU): a randomised, double-blind, placebo-controlled trial. Lancet Respir Med, 2013. 1(7): p. 515–23.

7. Mestres Gonzalvo, C., et al., Validation of an automated delirium prediction model (DElirium MOdel (DEMO)): an observational study. BMJ Open, 2017. 7(11): p. e016654.

8. Grover, S. and N. Kate, Assessment scales for delirium: A review. World J Psychiatry, 2012. 2(4): p. 58–70.

9. Terry, K.J., K.E. Anger, and P.M. Szumita, Prospective evaluation of inappropriate unable-to-assess CAM-ICU documentations of critically ill adult patients. J Intensive Care, 2015. 3: p. 52.

10. Thom, R.P., et al., A Comparison of Early, Late, and No Treatment of Intensive Care Unit Delirium With Antipsychotics: A Retrospective Cohort Study. Prim Care Companion CNS Disord, 2018. 20(6).

11. Knauert, M.P. and M.A. Pisani, Dexmedetomidine for hyperactive delirium: worth further study. J Thorac Dis, 2016. 8(9): p. E999-E1002.

12. Girard, T.D., et al., Haloperidol and Ziprasidone for Treatment of Delirium in Critical Illness. N Engl J Med, 2018. 379(26): p. 2506–2516.

13. Pluye, P. and Q.N. Hong, Combining the power of stories and the power of numbers: mixed methods research and mixed studies reviews. Annu Rev Public Health, 2014. 35: p. 29–45.

14. Moher, D., et al., Preferred reporting items for systematic reviews and meta-analyses: the PRISMA statement. Int J Surg, 2010. 8(5): p. 336–41.

15. Guo, Z., et al., Postoperative Delirium in Severely Burned Patients Undergoing Early Escharotomy: Incidence, Risk Factors, and Outcomes. J Burn Care Res, 2017. 38(1): p. e370-e376.

16. Kumar, A.K., et al., Delirium after Cardiac Surgery: A Pilot Study from a Single Tertiary Referral Center. Annals of Cardiac Anaesthesia, 2017. 20(1): p. 76–82.

17. Bilge, E.Ü., et al., The Incidence of Delirium at the Postoperative Intensive Care Unit in Adult Patients. Turkish Journal of Anaesthesiology and Reanimation, 2015. 43(4): p. 232–239.

18. Norkienė, I., et al., Incidence and Risk Factors of Early Delirium after Cardiac Surgery. BioMed Research International, 2013. 2013: p. 323491.

19. Saporito, A. and E. Sturini, Incidence of postoperative delirium is high even in a population without known risk factors. Journal of Anesthesia, 2014. 28(2): p. 198–201.

20. Visser, L., et al., Predicting postoperative delirium after vascular surgical procedures. Journal of Vascular Surgery, 2015. 62(1): p. 183–189.

21. Wesselink, E.M., et al.,Intraoperative hypotension and delirium after on-pump cardiac surgery. Br J Anaesth, 2015. 115(3): p. 427–33.

22. Hori, D., et al., Blood Pressure Deviations From Optimal Mean Arterial Pressure During Cardiac Surgery Measured With a Novel Monitor of Cerebral Blood Flow and Risk for Perioperative Delirium: A Pilot Study. Journal of cardiothoracic and vascular anesthesia, 2016. 30(3): p. 606–612.

23. Kim, M.Y., et al., DELirium Prediction Based on Hospital Information (Delphi) in General Surgery Patients. Medicine, 2016. 95(12): p. e3072.

24. Wang, S.-H., et al., Predisposing Risk Factors for Delirium in Living Donor Liver Transplantation Patients in Intensive Care Units. PLOS ONE, 2014. 9(5): p. e96676.

25. Yilmaz, S., et al., Dopamine Administration is a Risk Factor for Delirium in Patients Undergoing Coronary Artery Bypass Surgery. Heart, Lung and Circulation, 2016. 25(5): p. 493–498.

26. Zhang, W.-y., et al., Risk factors for postoperative delirium in patients after coronary artery bypass grafting: A prospective cohort study. Journal of Critical Care, 2015. 30(3): p. 606–612.

27. Liu, Z., et al., Incidence and Risk Factors of Delirium in Patients After Type-A Aortic Dissection Surgery. Journal of Cardiothoracic and Vascular Anesthesia, 2017. 31(6): p. 1996–1999.

28. Kazmierski, J., et al., Cortisol levels and neuropsychiatric diagnosis as markers of postoperative delirium: a prospective cohort study. Critical Care, 2013. 17(2): p. R38–R38.

29. Hori, D., et al., Arterial pressure above the upper cerebral autoregulation limit during cardiopulmonary bypass is associated with postoperative delirium. BJA: British Journal of Anaesthesia, 2014. 113(6): p. 1009–1017.

30. Choi, N.Y., et al., Development of a nomogram for predicting the probability of postoperative delirium in patients undergoing free flap reconstruction for head and neck cancer. European Journal of Surgical Oncology (EJSO), 2017. 43(4): p. 683–688.

31. Guo, Y., et al., Prevalence and risk factors of postoperative delirium in elderly hip fracture patients. Journal of International Medical Research, 2016. 44(2): p. 317–327.

32. Guenther, U., et al., Predisposing and precipitating factors of delirium after cardiac surgery: a prospective observational cohort study. Ann Surg, 2013. 257(6): p. 1160–7.

33. Ogawa, M., et al., Preoperative exercise capacity is associated with the prevalence of postoperative delirium in elective cardiac surgery. Aging Clinical and Experimental Research, 2017.

34. Parente, D., et al., Congestive heart failure as a determinant of postoperative delirium. Rev Port Cardiol, 2013. 32(9): p. 665–71.

35. Bakker, R.C., et al., Preoperative and operative predictors of delirium after cardiac surgery in elderly patients. European Journal of Cardio-Thoracic Surgery, 2012. 41(3): p. 544–549.

36. Serafim, R.B., et al., Delirium in postoperative nonventilated intensive care patients: risk factors and outcomes.Annals of Intensive Care, 2012. 2: p. 51–51.

37. Emma, L.R., et al., Epidemiology and outcomes of people with dementia, delirium, and unspecified cognitive impairment in the general hospital: prospective cohort study of 10,014 admissions. BMC Medicine, Vol 15, Iss 1, Pp 1–12 (2017), 2017(1): p. 1.

38. Lescot, T., et al., Postoperative delirium in the intensive care unit predicts worse outcomes in liver transplant recipients. Can J Gastroenterol, 2013. 27(4): p. 207–12.

39. Yoshitaka, S., et al., Perioperative plasma melatonin concentration in postoperative critically ill patients: its association with delirium. Journal Of Critical Care, 2013. 28(3): p. 236–242.

40. Lopez, M.G., et al., Intraoperative cerebral oxygenation, oxidative injury, and delirium following cardiac surgery. Free Radical Biology and Medicine, 2017. 103: p. 192–198.

41. Whitlock, E.L., et al., Postoperative Delirium in a Substudy of Cardiothoracic Surgical Patients in the BAG-RECALL Clinical Trial. Anesthesia and analgesia, 2014. 118(4): p. 809–817.

42. Silverstein, J.H., et al., Central nervous system dysfunction after noncardiac surgery and anesthesia in the elderly. Anesthesiology, 2007. 106(3): p. 622–8.

43. Vasunilashorn, S.M., et al., Cytokines and Postoperative Delirium in Older Patients Undergoing Major Elective Surgery. J Gerontol A Biol Sci Med Sci, 2015. 70(10): p. 1289–95.

